# (Poly)phenols in Human Breast Milk and their health benefits for the newborn

**DOI:** 10.1101/2023.03.27.23287781

**Authors:** Diogo Carregosa, Inês P. Silva, Carolina Teixeira, Mariana Baltazar, Rocio García-Villalba, Filipa Soares Vieira, Mónica Marçal, Madalena Tuna, Cláudia N. Santos

**Affiliations:** iNOVA4Health, NOVA Medical School | Faculdade de Ciências Médicas, Universidade NOVA de Lisboa, Campo dos Mártires da Pátria, 1169-056 Lisboa, Portugal; Laboratory of Food & Health, Research Group on Quality, Safety, and Bioactivity of Plant Foods, CEBAS-CSIC, Murcia, Spain; Hospital de São Francisco Xavier - Centro Hospitalar de Lisboa ocidental E.P.E; CHRC, NOVA Medical School | Faculdade de Ciências Médicas, Universidade NOVA de Lisboa, Campo dos Mártires da Pátria, 1169-056 Lisboa, Portugal

**Keywords:** Polyphenols, Breast Milk, Newborn, Metabolites, Concentrations

## Abstract

Human breast milk is the first food source available to newborns and is responsible for healthy growth and development during the first months of life. Human breast milk contains vitamins, hormones, cytokines, microbiota, and immune cells that are responsible for such healthy conditions. Nonetheless, knowledge of the vast array of molecules present in human breast milk and their potential health effects is still lacking. The effects of mothers’ diets on the molecules present in human breast milk are also generally unknown. The health benefits of (poly)phenols have been largely increasing but their presence in breast milk has been put largely aside. The impact of the mother’s diet in the presence and quantification of these molecules in milk has also been overlooked. Above all, the potential benefits of (poly)phenols for newborns are just a vast emptiness of what is known about (poly)phenol research.

## 1. Introduction

Human breast milk is produced by the mammary gland and is essential for the healthy growth and development of newborns. Human breast milk is characterized by a huge complexity and diversity of components, including nutrients, such as proteins, carbohydrates, and lipids (1); bioactive components, such as immunoglobulins, stem cells, cytokines, hormones, growth factors, and phytochemicals (2); and a vast array of bacteria, such as *Staphylococcus, Streptococcus, Bifidobacterium, Lactobacillus, and Clostridium* cluster XIVa–XIVb (3). This chemical and biological diversity of agents in its composition provides the infant with well-balanced nutrition and protection against potential infectious pathogens as the infant immune system develops (4). For these reasons and for optimal health and growth, the World Health Organization (5) recommends that infants be exclusively breastfed for the first six months of life, and that breastfeeding continues to be an important part of the diet until the infant is at least two years old. In addition to nutrients, vitamins, cells, and other compounds beneficial for health, it is not surprising that breast milk also contains some undesirable molecules, for example xenobiotics. Environmental xenobiotics in air, water and soil are absorbed by organisms through ingestion, inhalation or dermal absorption and enter the human food chain and often can be found in milk samples (6).

Human breast milk is a dynamic fluid that arises from the lactation process and develops to the different newborn needs. Breast milk can be classified into different stages based on its chemical composition. The first stage is pre-colostrum. This occurs as result of the process of lactogenesis that starts as early as the second trimester of pregnancy, the 16^th^ week of pregnancy, due to the increased levels of estrogen and progesterone in the circulation (7). From day 5 up until day 7 after birth, colostrum is secreted and has a sticky consistency of yellow color given by the presence of carotenes from dietary intake. Colostrum is characterized by its nutritional, immunological and growth functions. The third stage is the transition milk produced between days 4 and 15 after childbirth, which shows a white color due to the emulsification of fats and the strong presence of calcium caseinate (7). Finally, mature milk, is produced from the 15^th^ postpartum day looking like a thinner and diluted milk (8). During the different maturation stages of human breast milk, a high degree of interindividual differences can be expected in milk that may arise from differences in milk production and possible differences in transport of the molecules and other milk components like cells from blood to milk. Moreover, these differences are expected to be at least partially explained by a different dietary intake of the mother. However, we still lack knowledge on how diets like the Mediterranean diet, or vegetarian diets, rich in phytochemicals, might influence on the content of these compounds or their metabolites in human breast milk and how these might impact the health and development of the newborn.

Thousands of different dietary phytochemicals have been identified in human breast milk, of which carotenoids and (poly)phenols (9), represent by far the largest groups (10,11). In this study, we focused on the role of (poly)phenols such as phenolic acids, flavonoids, stilbenes, coumarins, lignans, and tannins (Figure 1) and their host and microbial metabolites for their well-documented protective and health-promoting properties, which are less understood compared to carotenoids (12).

**Figure 1.**
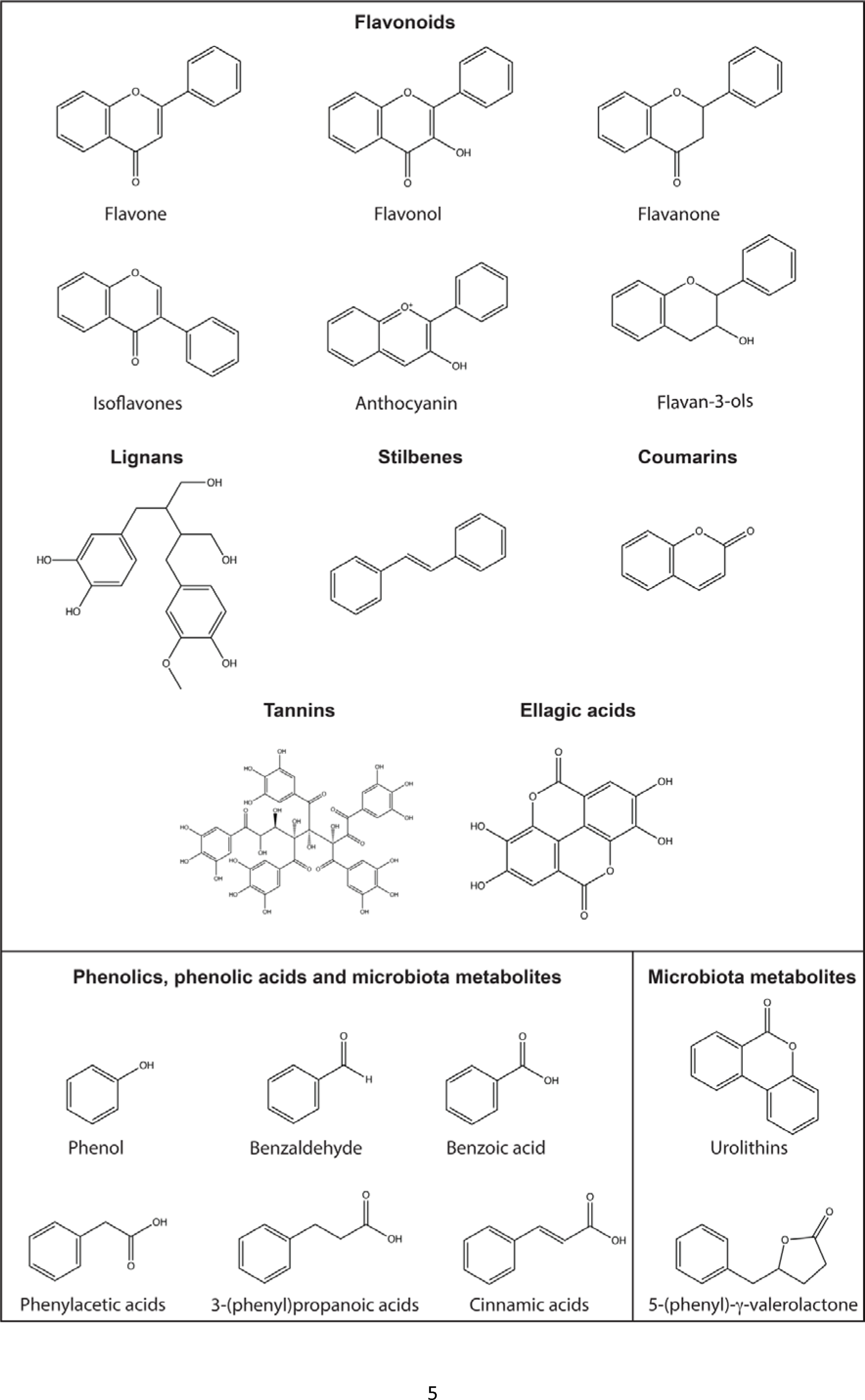
Chemical structures of common dietary (poly)phenols and microbiota metabolites. In the figure are represented common dietary (poly)phenols like: flavonoids, lignans, stilbenes, coumarins, tannins and ellagic acids. Furthermore, phenolics and phenolic acids are present in diet and can also result from the microbiota. Meanwhile, some molecules are only generated from the microbiota like urolithins from ellagic acids and 5-(phenyl)-γ-valerolactones from flavan-3-ols.

(Poly)phenols, a class of phytochemicals containing at least one phenolic ring, that are present in large quantities in fruits and vegetables like soybeans, cocoa beans, oranges and berries, as well as tea and coffee (13). They are believed to have health benefits, including protection against oxidative stress and inflammation, and may decrease the risk of chronic and degenerative diseases such as neurodegenerative disorders, obesity, cancer, and diabetes (14–18). They are classified in flavonoids and non-flavonoids where the latter include lignans, stilbenes, coumarins, tannins and simple phenolics/phenolic acids (Figure 1). Flavonoids represent the vast majority of (poly)phenols present in a fruit- and vegetable-rich diet, and include different subclasses such as: flavones, isoflavones, flavan-3-ols, flavonols, flavanones and anthocyanins. These (poly)phenols have demonstrated biological activities, including antioxidant and anti-inflammatory activities, consistent with the promotion of vascular health, bone health, and cognitive function (14,16,19). It has been demonstrated that the biological activities of all these phenolic compounds may be mediated by their metabolites which are produced *in vivo* (host and microbial metabolites), as in the case of urolithins, the ellagic acid microbial metabolites (20) and phenolic acids, the flavonoid microbial metabolites (16,21).

## 2. Absorption, metabolism, and distribution of (poly)phenols

Owing the wide variety of (poly)phenols, like flavonoids, phenolic acids, ellagic acids, among others, bioavailability varies a lot from one family to another. Some flavonoids like anthocyanins, isoflavones such as genistein and daidzein and the flavanol quercetin can be partially absorbed as early as in the stomach(22–25). However, most flavonoids are found in foods conjugated with sugars, esterified with organic acids, and polymerized, which decreases their absorption in the stomach, reaching the small intestine. In the intestine intact glycosides may undergo the action of lactase phloridzin hydrolase or cytosolic β-glucosidase breaking the sugar bond and releasing the respective aglycones which are subsequently absorbed. (Poly)phenols that are not absorbed in the small intestine reach the colon where they are transformed by the gut microbiota (Figure 2). In the case of flavonoids, they undergo catabolic reaction by the gut microbiota producing the low molecular weight phenolic metabolites also known by simple phenolics and phenolic acids, some of which common to those found in plants and already absorbed in initial phases of digestion, like benzoic acids (13,26). In fact, simple phenolics and phenolic acids present in the gut might result from two distinct pathways i) directly from the intake of fruits and vegetables or ii) the result of gut microbiota catabolism of flavonoids. Meanwhile, several (poly)phenols metabolites can only be produced by gut microbiota. This is the case of equol, produced exclusively by gut microbiota catabolism of the isoflavone daidzein, enterodiol and enterolactone, produced by catabolism of lignans, 5-(phenyl)-γ-valerolactones characteristic microbial metabolites of flavan-3-ols or urolithins the gut microbiota metabolites of ellagitannins and ellagic acid (27–29). In the case of equol and urolithins the presence of specific bacteria in the microbiota of the volunteers allowed to classify them in different metabotypes (20). For equol the two metabotypes differentiate equol-producers and equol-non producers. Meanwhile for urolithins three different urolithin metabotypes exist according to the capability of gut microbiota of the host to catabolize ellagic acid. These are metabotype A: producers of urolithin A; metabotype B: producers of urolithin A and also isourolithin A and/or urolithin B, and metabotype 0: non producers of urolithins (20,30).

**Figure 2.**
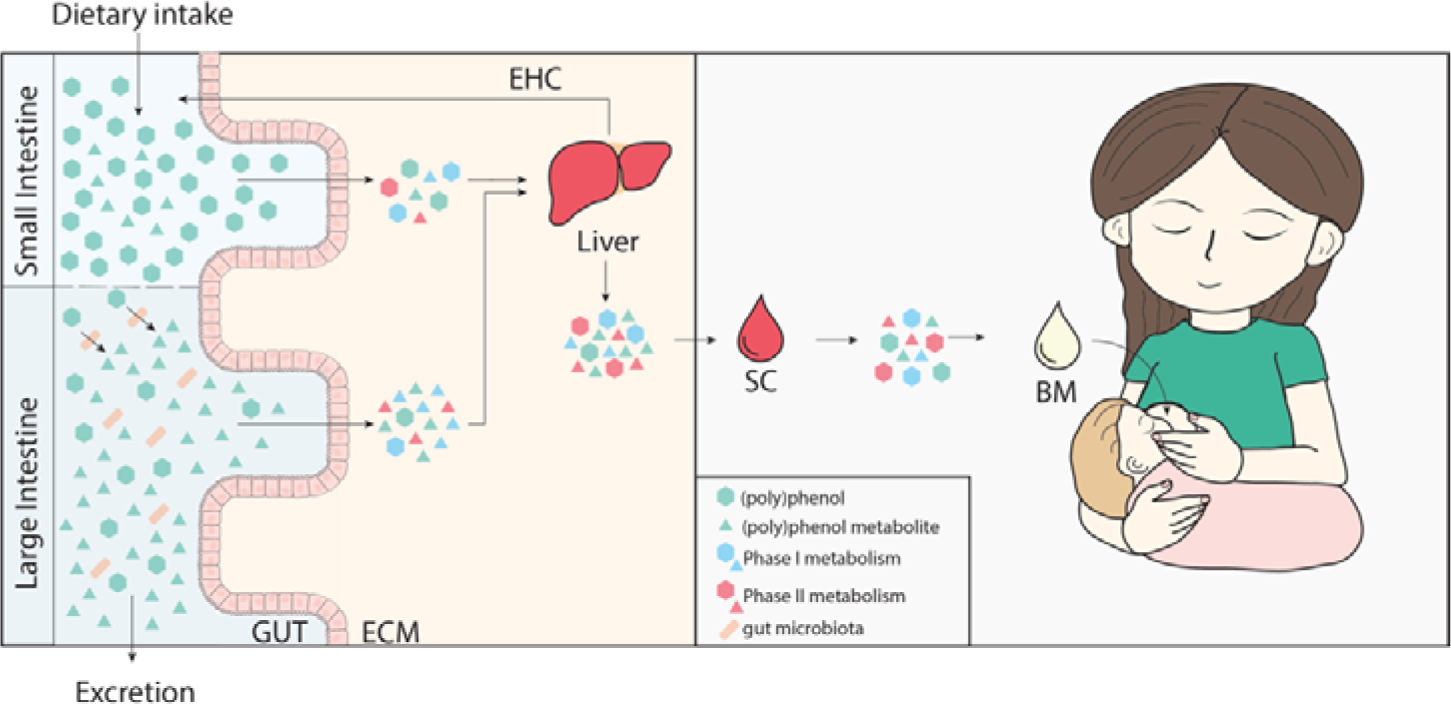
(Poly)phenol journey from dietary intake to breast milk in mothers and delivery the infant. (Poly)phenols are up taken from the diet reaching the gut. In the mother’s gut these (poly)phenols are absorbed and potentially modified by phase I and II metabolic reactions. Meanwhile (poly)phenols may be catabolized by gut microbiota into low molecular weight phenolics that are also up taken and undergo phase I and II metabolic reactions. All these compounds reach the liver and are released into circulation where they may reach breast milk. EHC – enterohepatic circulation; SC – systemic circulation; BM – breast milk.

Directly after their absorption in the small intestine or in the colon, (poly)phenols and their gut microbiota catabolites are extensively metabolized at both the intestinal and hepatic levels. Independently of the type of (poly)phenol, all undergo phase I and II metabolic reactions mediated by sulfotransferases, methyltransferases and glucuronyltransferases. Once in portal circulation, (poly)phenols metabolites travel into the liver where they undergo further phase I and II metabolic reactions. From the liver, (poly)phenols or their metabolites may reenter the intestine through bile salts and the enterohepatic circulation (EHC) or enter systemic circulation (Figure 2). In circulation, (poly)phenols metabolites spread across the different organs and tissues to exert their potential biological effects or are retrained in the kidneys and eliminated through urine. Importantly, several phenolic metabolites resulting from dietary intake or gut microbiota catabolism of flavonoids are also produced by some endogenous pathways. Some examples of molecules shared between pathways may include hippuric acid, homovanillic acid and 3′,4′-dihydroxyphenylacetic acid.

In lactating women, different (poly)phenols and their metabolites, mainly flavonoids, phenolic acids and ellagic acid and urolithin metabolites, have been detected in breast milk, as will be discussed ahead. These are expected to travel from blood circulation reaching the mammary glands (Figure 2), although the mechanism of transport has not been fully characterized. (Poly)phenols or their metabolites present in breast milk will start a new cycle of digestion as they will be ingested by the newborn infant. Importantly due to early stages of gut microbiota development in the infant, the amount of (poly)phenol metabolism expected in the infant is limited in comparison with the mother, but much about the infant microbiota capability to metabolize (poly)phenols is still unknown.

## 3. (Poly)phenolic content in Human Breast Milk

### 3.1 Search strategy

First, we searched the most well-known metabolomic and/or (poly)phenolic databases regarding presence of (poly)phenols in breast milk, e.g., HMDB, phenol-explorer, phyto-hub, metabolomics workbench, among others(31–33). Overall, only HMDB reported detection and quantification of (poly)phenols in human breast milk (34). A total of 122 metabolites have been detected in human breast milk according to HMDB, mainly including amino acids, short-chain-fatty acids, sugars, and carotenoids. From this list the only (poly)phenol metabolite reported was hippuric acid (34). Unfortunately, hippuric acid is also common to endogenous pathways and is difficult to pinpoint its origin (endogenous versus dietary).

Afterwards, we searched PubMed for articles containing the words: “human milk” or “human breast milk” and “polyphenol” or “flavonoid”. In total, we found 63 articles of which 2 were excluded due to duplication. Furthermore, 7 additional records were added since were found through other meanings (e.g., through in-text citing references). From the 68 records, 4 have been excluded since were not available in English. Furthermore, 45 articles have been excluded since they did not perform detection and/or quantification of (poly)phenols or human breast milk has not been used. (Figure 3).

**Figure 3.**
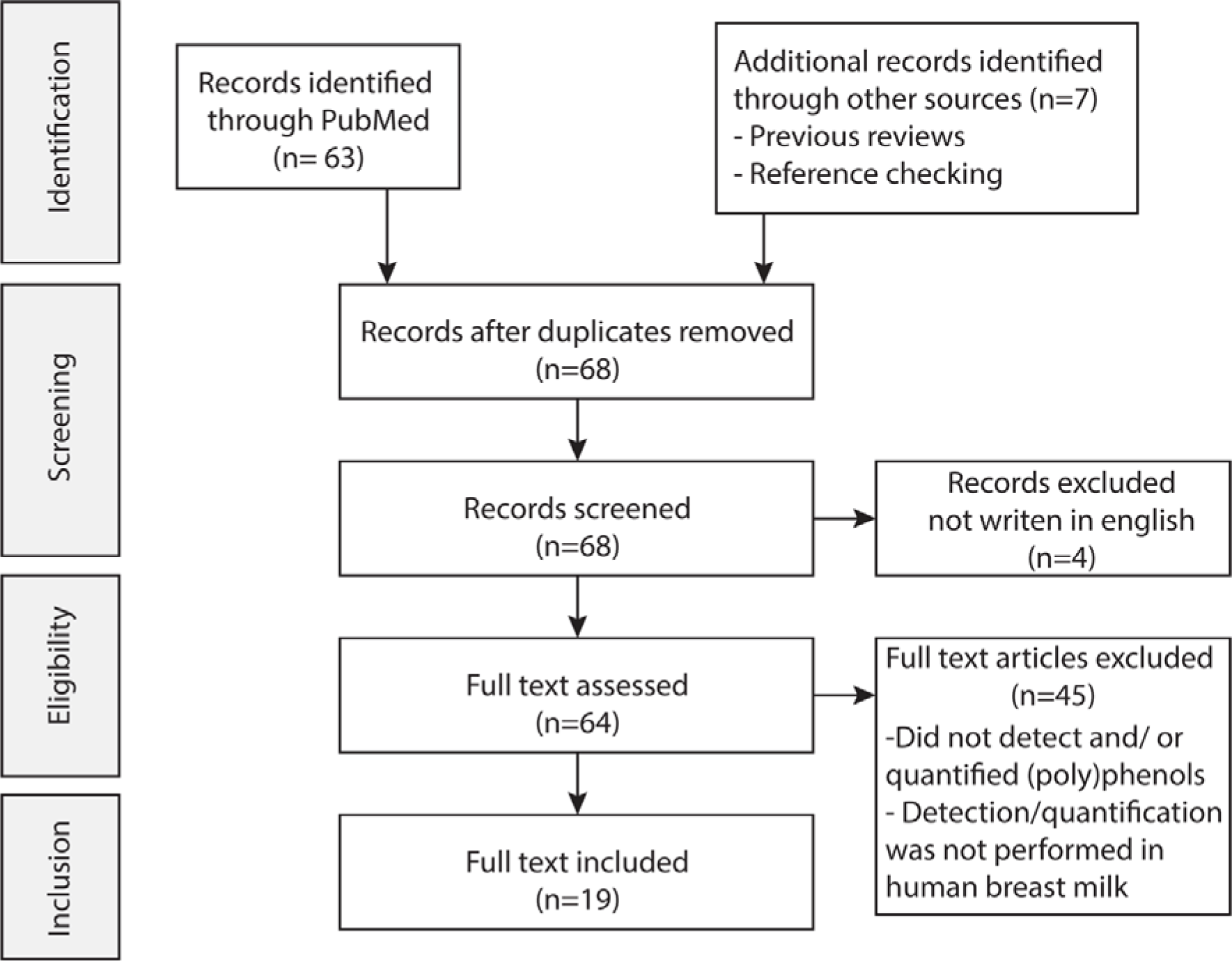
Search strategy for the literature review and revision process

### 3.2 Records reporting (poly)phenol quantification

From the list of records found, 19 have detected and quantified dietary (poly)phenols and/or metabolites in human breast milk. Of these, 12 have quantified (poly)phenols and/or metabolites from dietary intervention studies (Table 1) while 7 have quantified (poly)phenols and/or metabolites in free-living subjects (Table 2). Meanwhile, 15 of these 19 studies have quantified flavonoids, 4 have quantified ellagic acid or urolithins metabolites, and 6 studies have reported the quantification of low molecular weight phenolics (phenolic acids or simple phenols), see Table 1 and Table 2. Noteworthy 4 of the 7 studies quantifying (poly)phenols and/or metabolites in free-living subjects have included food-frequency questionnaires to the participants of the study to monitor their diet.

**Table 1.**
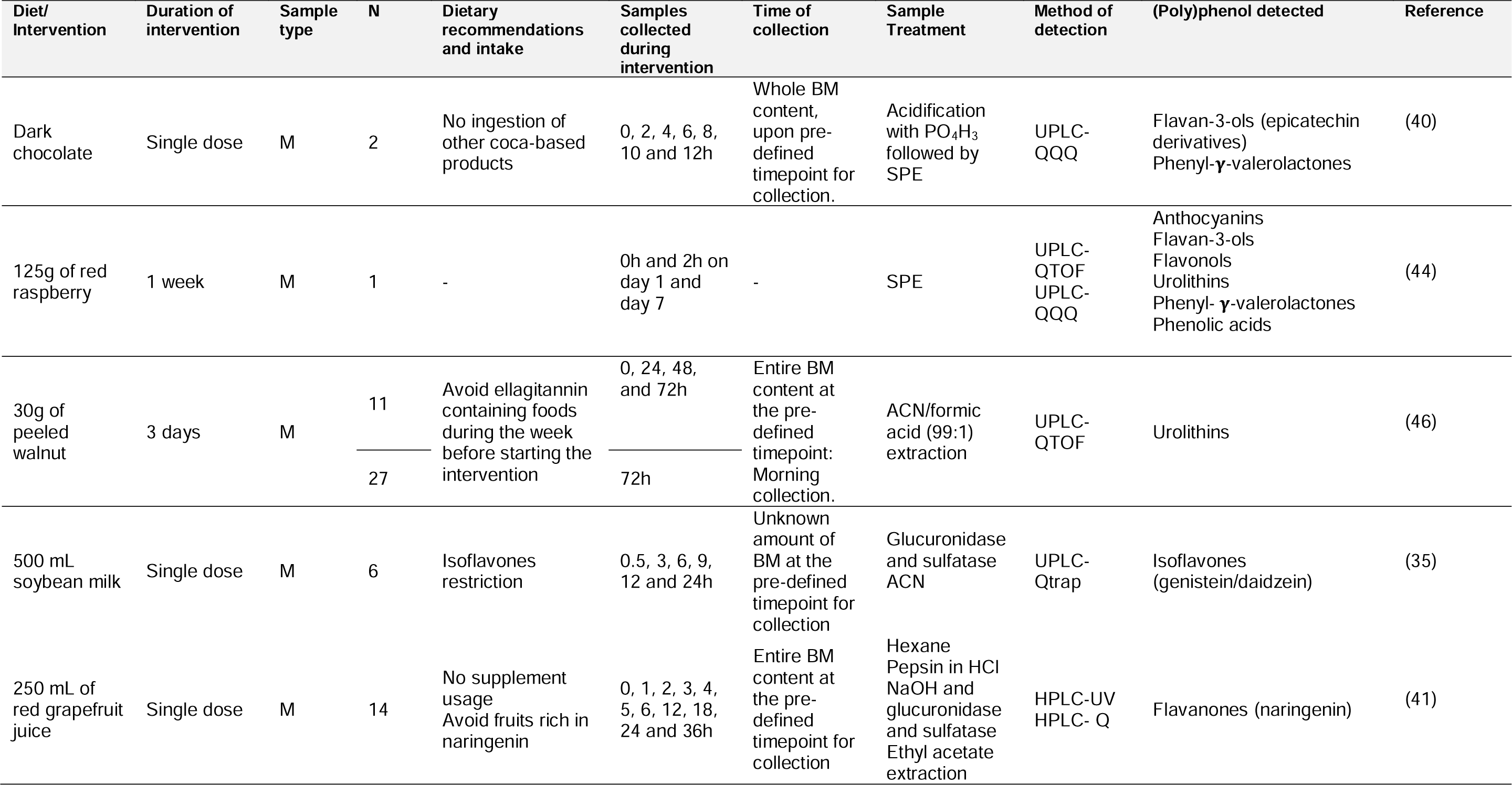
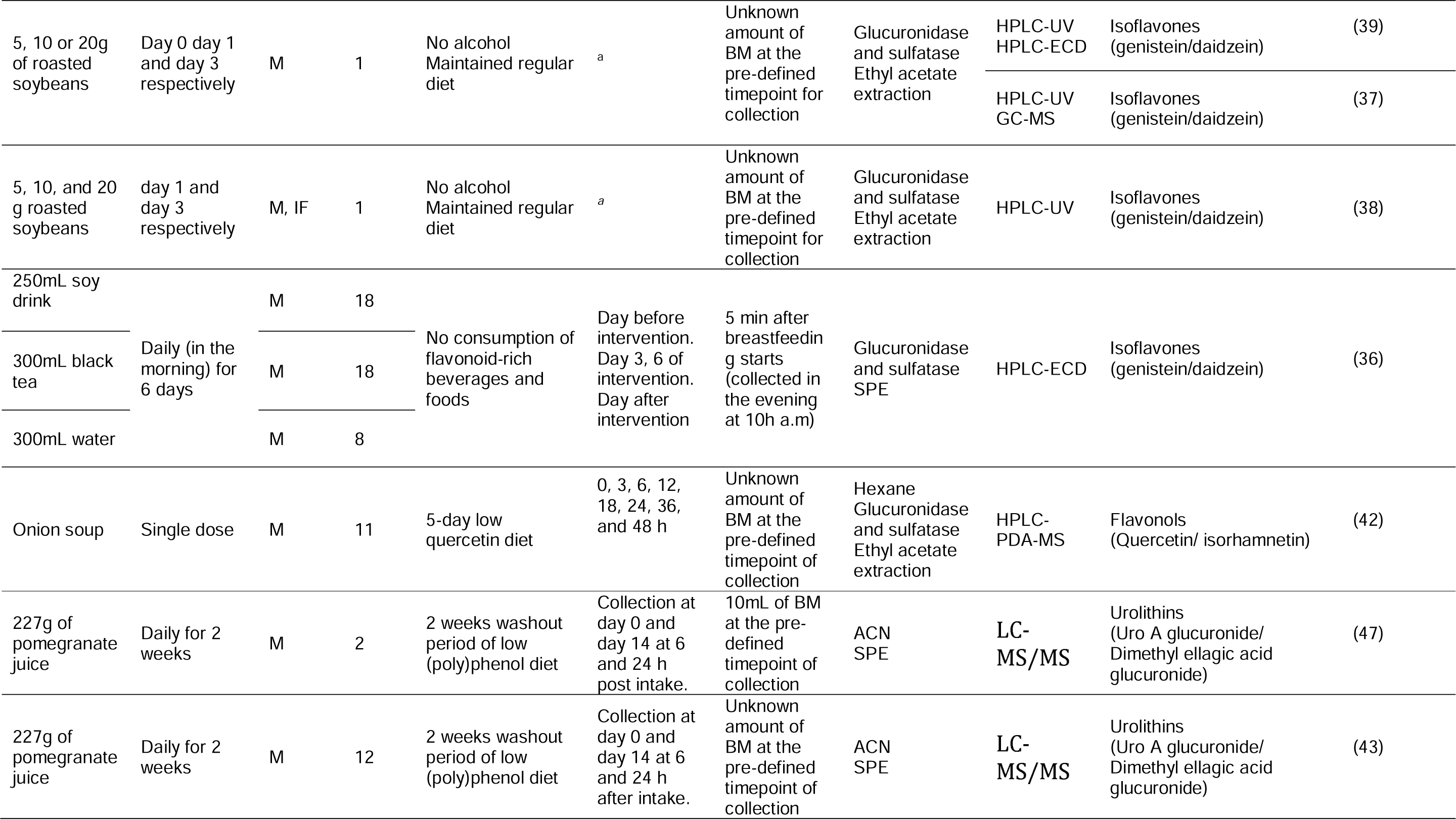
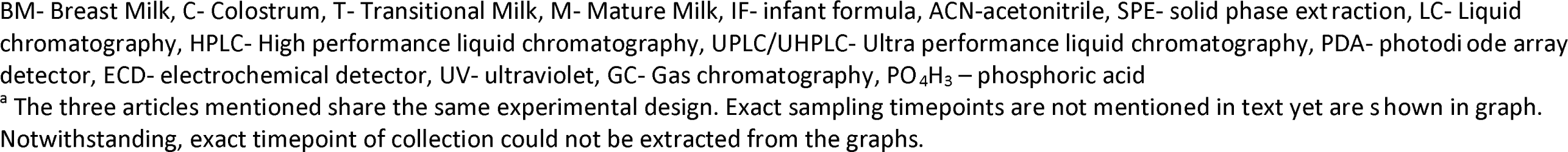
Characteristics of studies detecting and quantifying (poly)phenols in human breast milk samples of mothers undergoing dietary interventions.

**Table 2.**
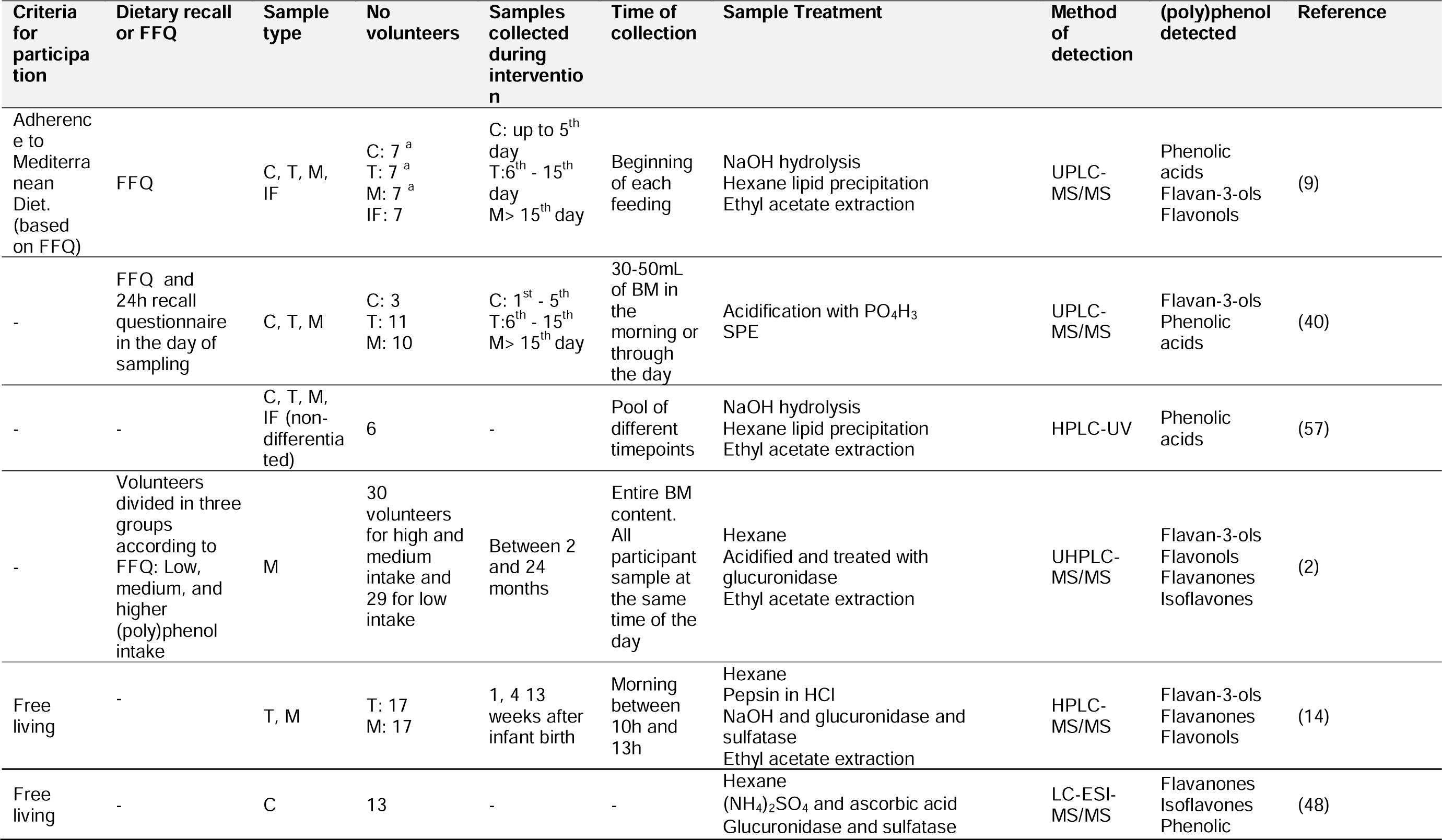
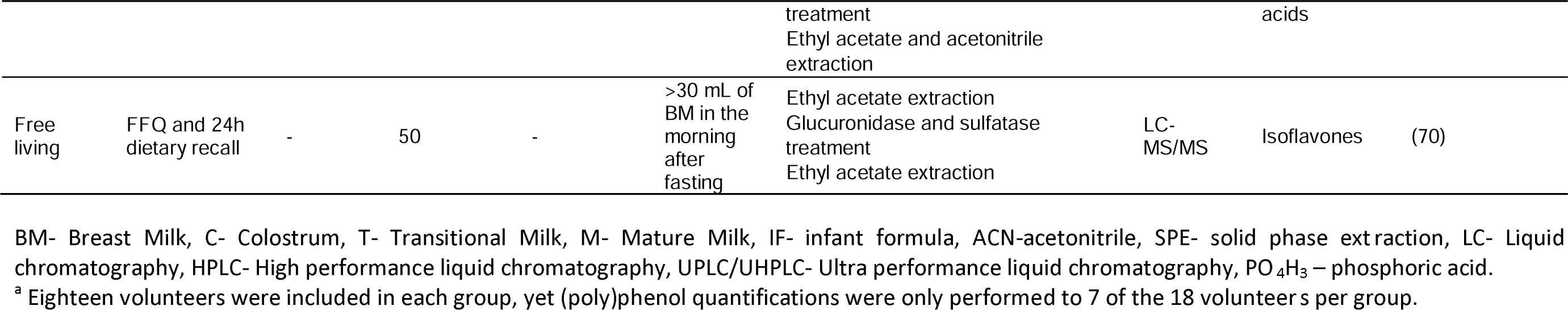
Characteristics of studies detecting and quantifying (poly)phenols in human breast milk samples of free-living mothers.

Several studies have shown the presence of isoflavones, mainly daidzein and genistein in human breast milk after dietary interventions using isoflavone rich foods like soybean milk (35,36), and roasted soybean (37–39). Meanwhile other flavonoids like flavan-3-ols and their microbial metabolites, 5-(phenyl)-γ-valerolactones were quantified in human breast milk after consumption of dark chocolate (40).Meanwhile, flavanones were detected after consumption of grapefruit juice (41), flavonols after onion soup intake (42) and the ellagitannin microbial metabolites, urolithins, after nuts and pomegranate consumption (20,43). In other studies, several (poly)phenol compounds, including different families of flavonoids, microbial metabolites, and phenolic acids, were tentatively identified in breast milk after consumption of red raspberry (44), and several in free-living conditions summarized in Table 2.

As previously described, breast milk composition evolves over time while the newborn matures. Most studies have only used matured milk as the sample to detect and quantify (poly)phenols and/or metabolites (Table 1 and Table 2). Meanwhile, transition milk has only been used in four studies and colostrum in three studies, all with free-living mothers. Notably, the three studies reporting (poly)phenol and/or metabolites quantification in colostrum have also used transitional milk and mature milk as samples.

Khymenets *et al.* was able to quantify some epicatechin phase II conjugates (epicatechin-4-glucuronide, epicatechin-2-sulfate and epicatechin-3-sulfate) and several gut microbiota derived metabolites like 5-(3′,4′-dihydroxyphenyl)-γ-valerolactone-sulfate and glucuronide conjugates (40). In this study, participants were in a free-living diet condition and answer to 24-hour dietary recall and food frequency questionnaires showing that the total (poly)phenol consumption through the diet did not significantly change during the different lactation periods. However, concentrations of the detected metabolites were very sporadic and varied significantly only being detected in a limited number of samples across the different lactation phases. For this reason, the authors were unable to compare the changes in concentrations amongst the different lactation stages.

Meanwhile, Sánchez-Hernández *et al.*, showed differences in the average amount of total (poly)phenols present in colostrum, transitional and matured milk, 661, 773 and 652 µg/100 mL respectively, of free-living individuals in comparison with infant formula, 375 µg/100 mL, (9). Noteworthy, this reflects in general on a 2-fold increase in total (poly)phenol content in human breast milk in comparison with formula milk (only 375 µg/100 mL), reflecting that formula milk does not recapitulate complete human breast milk concerning (poly)phenol content. Regarding the different stages of human breast milk, despite being a non-static fluid that changes with time, no difference in total (poly)phenol content or antioxidant capacity was observed among the three different types of milk.

In a different study Song *et al.*, was able to quantify several flavan-3-ols (epicatechin, epicatechin-gallate and epigallocatechin gallate), flavonols (quercetin and kaempferol) and flavanones (naringenin and hesperitin) in breast milk samples from free-living mothers at weeks 1, 4 and 13 postpartum (14). The concentrations of the those (poly)phenols did not seem to change, independently of the timepoint of collection (week 1, 4 or 13), with the only exception being kaempferol that significantly increased at week 13. The authors point out that the levels of flavonoids during the three periods suggest that (poly)phenols in human milk are generally not affected by lactation stage. Unfortunately, in this study the food intake of the mothers was not evaluated and thus the estimation of (poly)phenol intake for each mother was not performed. For this reason, it is possible that some of the variability in kaempferol concentrations might be due to the food intake of the mother. In fact, from the list of studies identified, the study by Lu *et al.* was the only that fully characterized and stratified samples according to the food intake (2). For that maternal intake of plant-based foods and (poly)phenols was estimated through 3-day dietary records and the use of Phenol-Explorer database. The study concluded that the intake of plant-based foods significantly affected phenolic composition in breast milk.

Meanwhile Romasko *et al.* showed that the amount of naringenin in breast milk in free living mothers with their everyday diet, resulted in an average naringenin concentration in breast milk of 823.24 nmol/L (41). Upon consumption of red grapefruit juice, rich in naringenin and naringin (naringenin-7-glucoside) this concentration did not significantly increase at 4 or 12 hours, 908.25 nmol/L ± 676.84 nmol/L, and 868.96 nmol/l ± 665.54 nmol/L, respectively. In this case, the consumption of a glass of grapefruit juice did not significantly affect naringenin levels in milk. Eventually the concentrations of naringenin decreased after 48 hours to 673.89 nmol/L. Meanwhile in a study by Jochum *et al.* two different interventions were used and compared against a control (300 mL water) drink: i) 250 mL of soy drink or ii) 300mL of decaffeinated black tea (36). Unfortunately, no flavonol was detected before or after the intervention with black tea. Yet, in the soy drink group, genistein and daidzein were detected, but only after the intervention.

The changes in values across these studies may be related to several factors. One of the most important is dietary exposure to different amounts of (poly)phenols and/or foods containing different/specific (poly)phenols. Dissimilarities in the dietary habits among different populations could lead to high variability in the results obtained. It is very important to define dietary habits during lactation to fully understand how these changes in (poly)phenol and/or metabolites quantification is dependent on the mother’s diet. However, there is still limited information on the relationship between maternal dietary intake and phenolic content in breast milk. Besides, although the concentration of the compounds is dependent on the dose ingested by the mother, other regular components of the matrix such as proteins, lipids and carbohydrates could affect the extraction and detection of these compounds.

Other important factors that could influence the detection and quantification of (poly)phenols and/or metabolites in human breast milk could be the kinetic of deposition in milk, interindividual differences in the metabolism and genetic/physiological factors and/or stability of (poly)phenols and/or metabolites in breast milk matrix. Although with the available studies, the time of milk collection has not been demonstrated to be a crucial factor, more studies with large population are needed to confirm these results. The methodology for the collection of breast milk samples could also be another critical point. Most of the time the volume of the milk produced by the mammary glands of the mothers is not measured, so the data obtained from the milk analysis only provide concentrations and not the total amount produced, what makes more difficult to compare among studies. Number of volunteers across the 19 studies herein reported has also shown a significant pitfall on the data available to researchers and clinicians. Until now only one study has used a sample size equal to 50 volunteers, while most studies have used sample sizes below 30 volunteers, with a significant number of studies with a sample size below 10 and some proof-of-concept/case studies with sample sizes of just one or two individuals. Such sample size, together with the normal interindividual variability detected in (poly)phenol bioavailability (26,45) clearly makes it difficult to understand the dynamics of (poly)phenols metabolites in breast milk. Larger populations are necessary to obtain final conclusions about concentrations of metabolites in breast milk and factors affecting.

Differences in the methodologies used for the extraction and analysis (identification and quantification) of these compounds in breast milk also hamper the comparison among studies.

The complexity of the milk matrix partly explains why the determination of (poly)phenol-derived metabolites in breast milk has hardly been investigated. Most of the methodologies are not optimized and validated and the recovery percentages of (poly)phenol metabolites are usually unknown for the different extraction protocols. Most previous studies have determined (poly)phenols in human milk after enzymatic hydrolysis (β-glucuronidase and sulfatase) and subsequent extraction with ethyl acetate (see Table 1 and Table 2). The enzymatic hydrolysis simplifies the separation and quantification of the major compounds and provides an idea of the total amount of a specific aglycone (free compound plus conjugated) but information about the nature of the metabolites available and the amount of individual conjugated metabolites is lost. Only 5 studies provide information about naturally occurring metabolites (glucuronides and sulfates) present in breast milk (40,43,44,46,47). Protein precipitation with acetonitrile or methanol and solid phase extraction (SPE) were also applied for the extraction of these compounds with and without previous enzymatic hydrolysis. In some cases, a previous clean up with hexane was applied to defat the samples and other methods to remove proteins were also used (hydrolysis with pepsin, salting-out assisted liquid-liquid extraction, precipitation with organic solvents, etc.). However, little information about recoveries and validation parameters was provided to evaluate the results and identify the best extraction protocol for each family of compounds. Only two studies have shown a comprehensive test between the different extraction methodologies (46,48). The data showed that acetonitrile: formic acid (95:1) extraction/protein precipitation method was the most effective method to obtain ellagic acid metabolites in comparison to methanol, hexane and SPE (46). Meanwhile Edyta Nalewajko-Sieliwoniuk showed that hexane defat followed by acidification and ethyl acetate: acetonitrile extraction was the most effective method for the extraction of several flavonoids and phenolic acids (48). Unfortunately, this protocol includes the use of β-glucuronidase and sulfatases. In general, it seems that ethyl acetate, which is frequently used for isolation of dietary phenolic compounds, is a good solvent for the extraction of aglycones after enzymatic hydrolysis. However, when more polar compounds such as glucuronide and sulfate conjugates are extracted, the recoveries are low and other protocols based of precipitation of protein with organic solvents or solid phase extraction should be used.

Another question that could affect the extraction and analysis of compounds in the different studies is the nature of the milk (content of protein and lipids) that could affect the extraction protocol with the formation of gel aggregates or the precipitation of some compounds.

Besides, the way of quantification is another important question that could influence the results in the different studies. In most cases the absence of commercial standards forces to quantify using other available standards, obtaining inaccurate results not comparable among studies.

### 3.3 Detected and quantified (poly)phenols in human breast milk

Across the 19 records reporting (poly)phenols and metabolites in human breast milk a total of 25 flavonoids, 58 low molecular weight phenolics and 9 ellagic acid metabolites were detected (Figure 4, Table 3-5). The list of flavonoids included the most representative subfamilies of flavonoids: anthocyanins, flavonols, flavan-3-ols, isoflavones and flavanones (Table 3, Figure 4). Interestingly besides sulfate and glucuronide conjugates some flavonoid glycoside conjugates have been found in human breast milk (Table 3). From the 9 ellagic acid metabolites, urolithin A, isourolithin A and urolithin B and respective sulfate and glucuronide metabolites were reported together with dimethyl ellagic acid-glucuronide, but not ellagic acid (Table 4, Figure 4). Regarding the low molecular weight phenolics detected, these ranged from benzoic acids, benzaldehydes, phenylacetic acids, 3-(phenyl)propanoic acids, cinnamic acids, 5-(phenyl)-γ-valerolactones, hippuric acids and chlorogenic acid (Table 5, Figure 4). Anthocyanins, isoflavones, flavanones and ellagic acid metabolites have not been detected in all types of milk, by contrast with the remaining (poly)phenolic classes that have been detected across all the different types of milk (colostrum, transitional and mature). It is however important to note that most studies have used one type of milk only, mature milk, thus explaining why several (poly)phenols have been detected in only one milk type.

**Figure 4.**
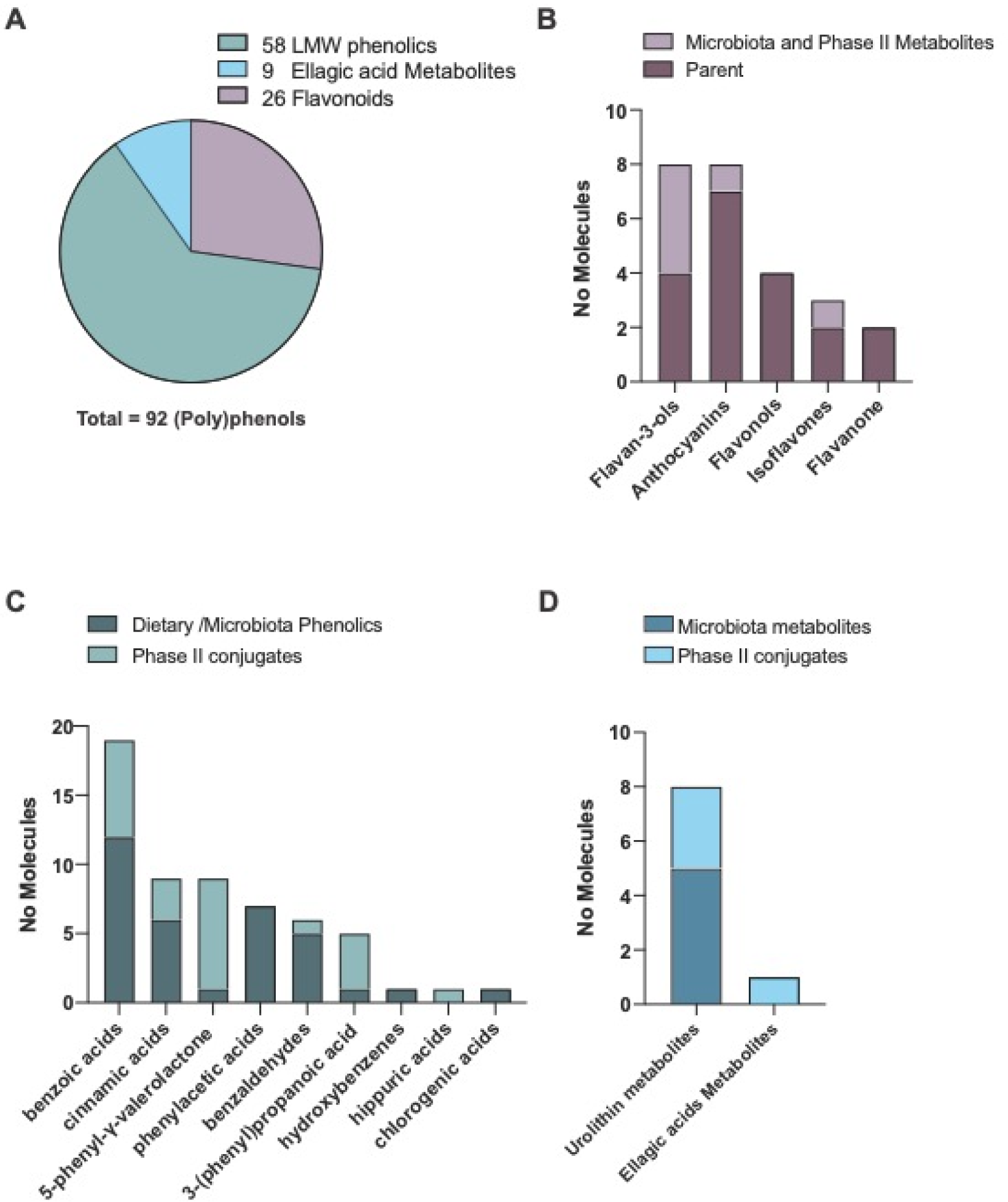
Ninety-two (poly)phenols and metabolites have been identified and/or quantified in human breast milk samples across the 19 reports identified. (A) From the overall list of compounds 58 were low molecular weight (LMW) phenolics, 9 ellagic acid metabolites, and 25 flavonoids. (B) The 25 flavonoids spread across flavan-3-ols, anthocyanins, flavonols, isoflavones and flavanones. (C) The low molecular weight phenolics and their phase II conjugates are divided into benzoic acids, cinnamic acids, 5-phenyl-γ-valerolactones, phenylacetic acids, benzaldehydes, 3-(phenyl)propanoic acids, hydroxybenzenes, hippuric acids and chlorogenic acid. (D) The 9 ellagic acids and/or metabolites and their phase II conjugates are divided into 8 urolithin metabolites and 1 ellagic acid metabolites. Individual concentrations for each (poly)phenol are reported in tables 3, 4, and 5.

**Table 3.**
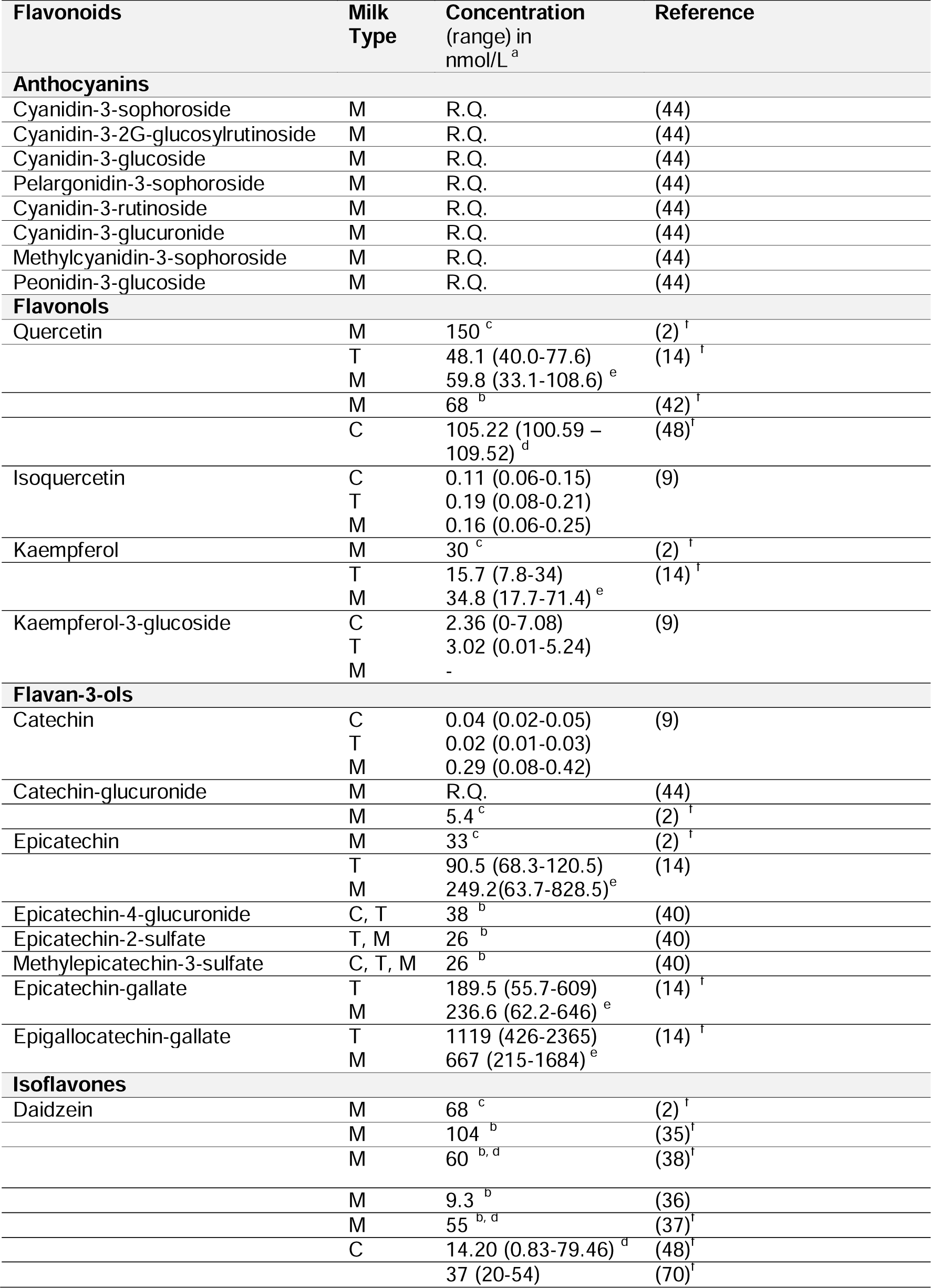
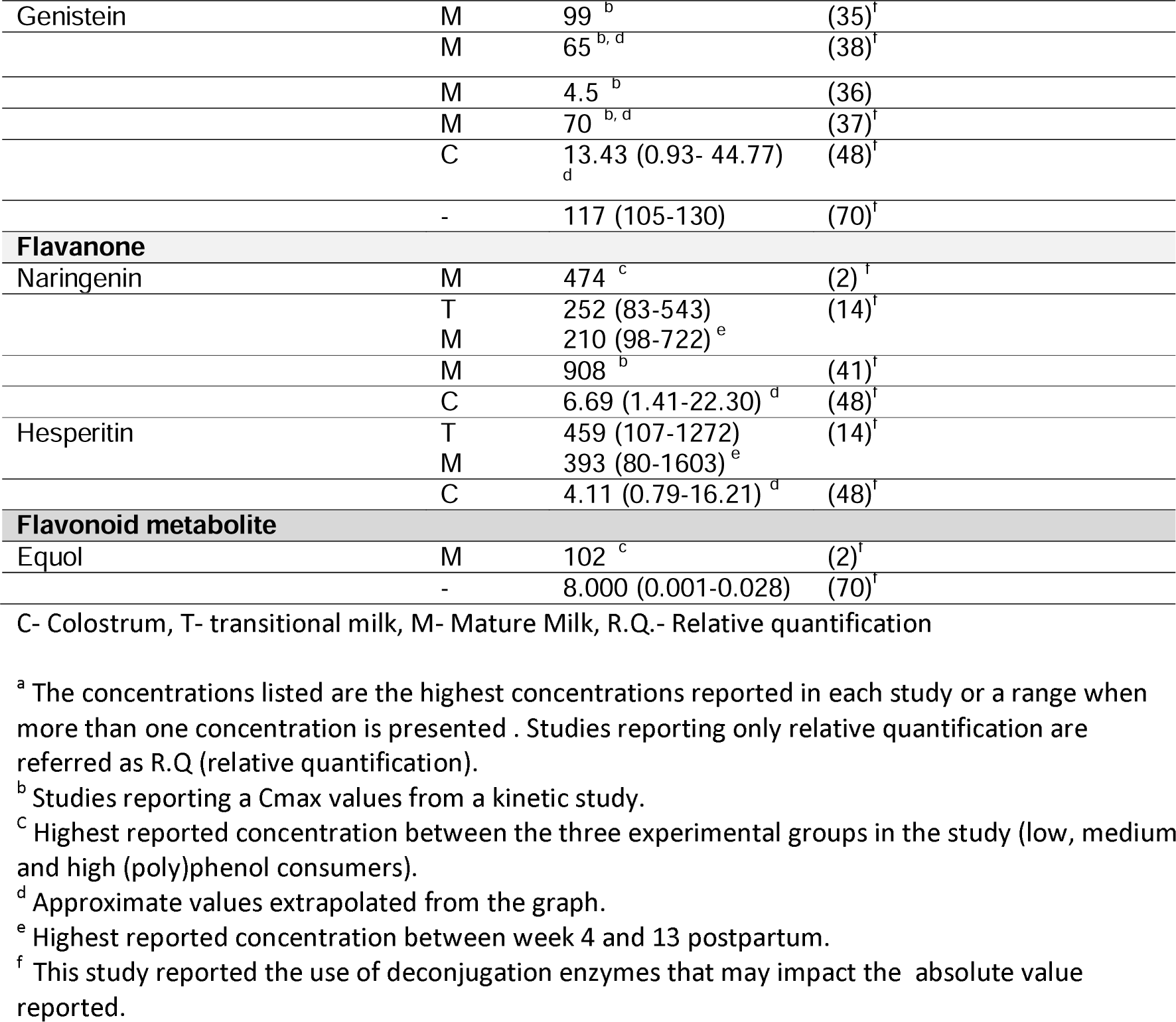
Dietary Flavonoids and metabolites detected in human breast milk, and their concentrations.

**Table 4.**
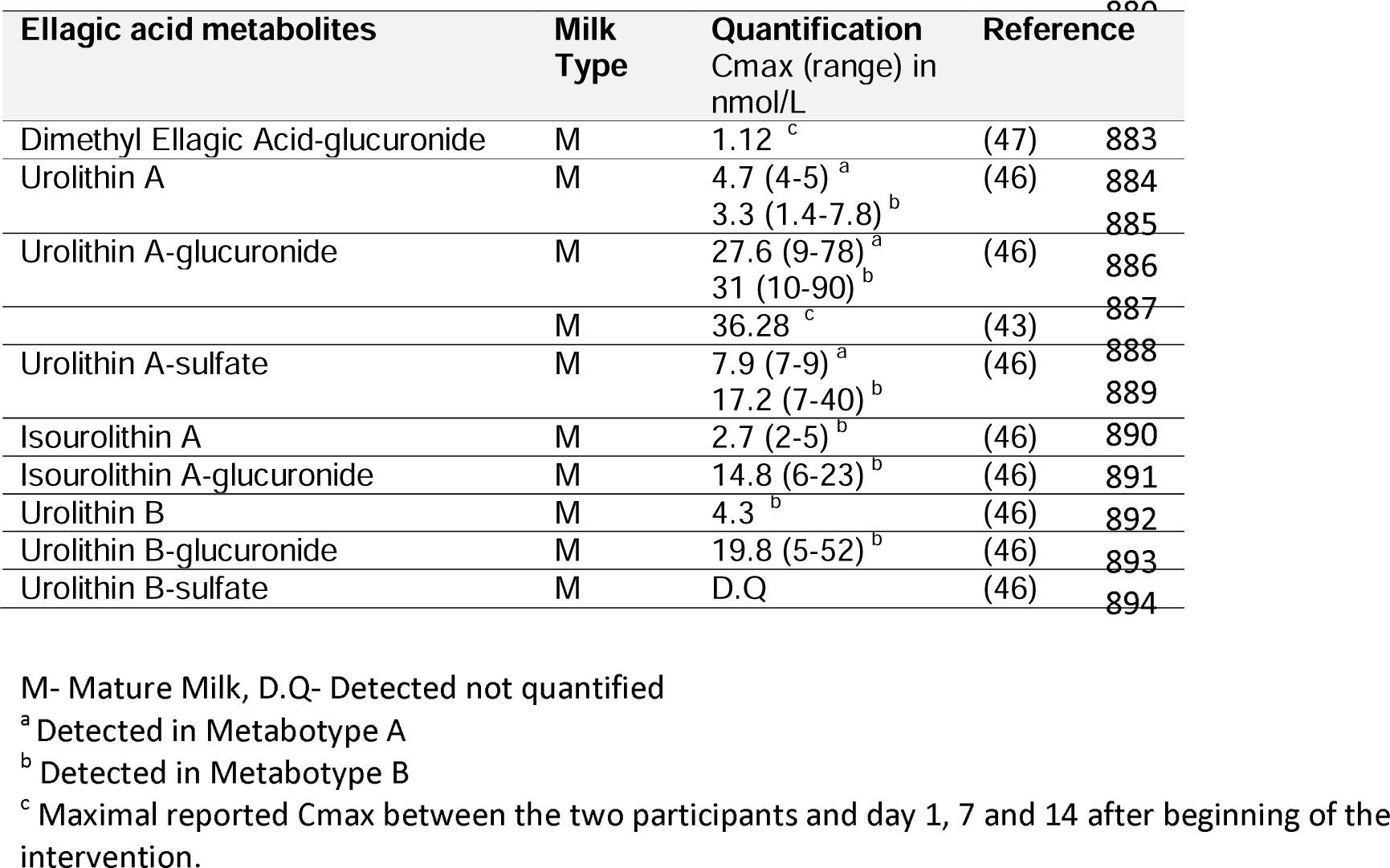
Ellagic acid metabolites detected and quantified in human breast milk, and their concentrations.

**Table 5.** Low molecular weight phenolics detected in human breast milk, and their concentrations.

| Phenolic Metabolite | Milk Type | Concentration (range) in nmol/L <sup>a</sup> | Reference |
| --- | --- | --- | --- |
| Benzenes |  |  |  |
| 1,2,3-trihydroxybenzene (pyrogallol) | C | 0.75 (0.34-1.73) | (9) |
|  | T | 0.95 (0.28-2.13) |  |
|  | M | 1.66 (1.41-3.91) |  |
| Benzoic acids |  |  |  |
| 3,4,5-trihydroxybenzoic acid (gallic acid) | C | 582 (375–721) | (9) |
|  | T | 661 (434–816) |  |
|  | M | 575 (434–816) |  |
|  | M | 61 <sup>c</sup> | (2) <sup>f</sup> |
|  | M | R.Q. | (44) |
|  | C | 186.92 (178.70-194.57) <sup>d</sup> | (48) <sup>f</sup> |
| 2,3-dihydroxybenzoic acid | M | R.Q. | (44) |
| 2,5-dihydroxybenzoic acid | C | 5.61 (4-8) | (9) <sup>f</sup> |
|  | T | 7.41 (3-15) |  |
|  | M | 6.43 (0.29-11) |  |
| 2,6-dihydroxybenzoic acid |  | R.Q. | (44) |
| 4-hydroxy-3-methoxybenzoic acid (vanillic acid) | C | 5.96 (6-7) | (9) <sup>f</sup> |
|  | T | 7.05 (6-8) |  |
|  | M | 6.56 (4-10) |  |
|  | M | R.Q. | (44) |
| 3-methoxybenzoic acid-4-sulfate (vanillic acid-sulfate) | M | R.Q. | (44) |
| 3-methoxybenzoic acid-4-glucuronide (vanillic acid-glucuronide) | M | R.Q. | (44) |
| 4-methoxy-3-hydroxybenzoic acid (Isovanillic acid) | M | R.Q. | (44) |
| 4-methoxy-benzoic acid-3-sulfate (Isovanillic acid-sulfate) | M | R.Q. | (44) |
| 3,4-dihydroxybenzoic acid (protocatechuic acid) | C | 15.16 (5-31) | (9) <sup>f</sup> |
|  | T | 22.55 (9-27) |  |
|  | M | 13.16 (3-20) |  |
|  | M | 770 <sup>c</sup> | (2) <sup>f</sup> |
|  | M | R.Q. | (44) |
| 3,4-dihydroxybenzoic acid-sulfate <sup>b</sup> (protocatechuic acid-sulfate) | M | R.Q. | (44) |
| 2,4,6-trihydroxybenzoic acid | C | 2.71 (1-6) | (9) |
|  | T | 2.68 (1-4) |  |
|  | M | 3.08 (0.25-6) |  |
| 4-hydroxybenzoic acid | C | 0.54 (0.24-0.90) | (9) |
|  | T | 0.67 (0.44-0.94) |  |
|  | M | 0.72 (0.26-1.9) |  |
|  | M | R.Q. | (44) |
|  | C, T, M | (4445-4598) <sup>d</sup> | (57) |
| benzoic acid-4-glucuronide | M | R.Q. | (44) |
| benzoic acid-4-sulfate | M | R.Q. | (44) |
| 3-hydroxybenzoic acid | C | 7.66 (3-14) | (9) |
|  | T | 9.94 (6-12) |  |
|  | M | 6.85 (3-8) |  |
| benzoic acid-3-glucuronide | M | R.Q. | (44) |
| methyl 4-hydroxybenzoate | M | R.Q. | (44) |
| methyl 3,4-dihydroxybenzoate | M | R.Q. | (44) |

| **Benzaldehydes** |  |  |  |
| --- | --- | --- | --- |
| 3,4-dihydroxybenzaldehyde<br>(protocatechualdehyde) | C | 9.63 (3-17) | (9) |
|  | T | 12.56 (7-16) |  |
|  | M | 7.46 (3-10) |  |
|  | M | R.Q. | (44) |
| 4-hydroxybenzaldehyde<br>(p-hydroxybenzaldehyde) | M | R.Q. | (44) |
|  | C | 1.09 (1-6) | (9) |
|  | T | 3.54 (1-5) |  |
|  | M | 1.62 (0.41-3) |  |
| benzaldehyde-4-glucuronide<br>(4-hydroxybenzaldehyde-glucuronide) | M | R.Q. | (44) |
| 4-hydroxy-3-methoxybenzaldehyde<br>(vanillin) | C | 5.84 (3-9) | (9) |
|  | T | 7.83 (7-10) |  |
|  | M | 5.56 (2-10) |  |
| 2-hydroxy-3-methoxybenzaldehyde<br>(o-vanillin) | C | 1.61 (1.41-1.62) | (9) |
|  | T | 6.06 (6-7) |  |
|  | M | 12.22 (0.30-32) |  |
| 2,4,6-trihydroxybenzaldehyde<br>(phloroglucinaldehyde) | M | R.Q. | (44) |
| **Cinnamic Acids** |  |  |  |
| cinnamic acid<br>(coumaric acid) | M | R.Q. | (44) |
| 4'-hydroxy-3'-methoxycinnamic acid<br>(ferulic acid) | C | 0.62 (0.06-1.23) | (9) |
|  | T | 0.71 (0.12-1.53) |  |
|  | M | 0.34 (0.14-0.57) |  |
|  | M | 129<sup>c</sup> | (2)<sup>i</sup> |
|  | M | R.Q. | (44) |
|  | C, T, M | (7339-7673)<sup>d</sup> | (57) |
| 3'-methoxycinnamic acid-4'-glucuronide<br>(ferulic acid-glucuronide) | M | R.Q. | (44) |
| 3',4'-dihydroxycinnamic acid<br>(caffeic acid) | C | 14.15 (4-17) | (9) |
|  | T | 17.28 (4-30) |  |
|  | M | 12.32 (1-24) |  |
|  | M | 188<sup>c</sup> | (2)<sup>i</sup> |
|  | C | 301.95 (129.32- 654.97)<sup>d</sup> | (48)<sup>i</sup> |
| 3',4'-dihydroxycinnamic acid-sulfate<sup>b</sup><br>(caffeic acid-sulfate) | M | R.Q. | (44) |
| 4'-methoxycinnamic acid | C | 0.24 (0.02-0.67) | (9) |
|  | T | 0.30 (0.14-0.48) |  |
|  | M | 0.91 (0.06-2.42) |  |
| 3',4'-dimethoxycinnamic acid<br>(dimethylcinnamic acid) | C | 0.14 (0.13-0.28) | (9) |
|  | T | 0.56 (0.15-0.25) |  |
|  | M | 0.18 (0.13-0.24) |  |
| 4'-hydroxycinnamic acid<br>(p-coumaric acid) | C | 0.44 (0.24-0.59) | (9) |
|  | T | 1.20 (0.27-2.31) |  |
|  | M | 0.29 (0.15-0.49) |  |
|  | C, T, M | (8474-8797)<sup>d</sup> | (57) |
| cinnamic acid-4'-sulfate<br>(p-coumaric acid-sulfate) | M | R.Q. | (44) |
| **3-(phenyl)propanoic acids** |  |  |  |
| 3-(3'-methoxyphenyl)propanoic acid-4'-glucuronide<br>(dihydroferulic acid-glucuronide) | M | R.Q. | (44) |
| 3'-methoxycinnamic acid-4'-sulfate<br>(dihydroferulic acid-sulfate) | M | R.Q. | (44) |
| 3-(3',4'-dihydroxyphenyl)propanoic acid-sulfate <sup>b</sup><br>(dihydrocaffeic acid-sulfate) | M | R.Q. | (44) |
| 3-(3'-hydroxyphenyl)propanoic acid | M | R.Q. | (44) |
| 3-(phenyl)propanoic acid-3'-sulfate<br>(3-(3-hydroxyphenyl)propanoic acid-sulfate) | M | R.Q. | (44) |
| **Phenylacetic acids** |  |  |  |
| 2'-hydroxyphenylacetic acid | M | R.Q. | (44) |
| 3'-hydroxyphenylacetic acid | M | R.Q. | (44) |
| 4'-hydroxyphenylacetic acid | M | R.Q. | (44) |
| 4'-hydroxyphenylacetic acid<br>(*p*-hydroxyphenylacetic acid) | C<br>T<br>M | 0.27 (0.05-0.53)<br>0.41 (0.35-0.52)<br>0.40 (0.07-0.93) | (9) |
| 3',4'-dihydroxyphenylacetic acid<br>(dopac) | M | 277 <sup>c</sup> | (2)<sup>f</sup> |
|  | M | R.Q. | (44) |
| 4'-hydroxy-3'-methoxyphenylacetic acid<br>(homovanillic acid) | M | R.Q. | (44) |
| 3'-hydroxy-4'-methoxyphenylacetic acid<br>(Isohomovanillic acid) | M | R.Q. | (44) |
| **5-(phenyl)-γ-valerolactone** |  |  |  |
| 5-(3',4'-dihydroxyphenyl)-γ-valerolactone-glucuronide (isomer 1) <sup>b</sup> | M | 26 | (40) |
| 5-(3',4'-dihydroxyphenyl)-γ-valerolactone-glucuronide (isomer 2) <sup>b</sup> | C<br>T<br>M | 37<br>(22-39)<br>(19-39) | (40) |
| 5-(3',4'-dihydroxyphenyl)-γ-valerolactone-sulfate | C<br>T<br>M | (165-781)<br>(24-1909)<br>(20-435) | (40) |
| 5-(3'-methoxyphenyl)-γ-valerolactone-4'-glucuronide | M | 24 | (40) |
| 5-(3'-methoxy-4'-hydroxyphenyl)-γ-valerolactone-sulfate <sup>e</sup> | T<br>M | (14-26)<br>(17-20) | (40) |
| 5-(3'-methoxy-4'-hydroxyphenyl)-γ-valerolactone-sulfate <sup>e</sup> | C<br>T<br>M | 20<br>20<br>17 (16.9-17.2) | (40) |
| 5-(3',4',5'-trihydroxyphenyl)-γ-valerolactone | M | R.Q. | (44) |
| 5-(4'-hydroxyphenyl)-γ-valerolactone-3'-glucuronide | M | R.Q. | (44) |
| 5-(3',4'-dihydroxyphenyl)-γ-valerolactone-sulfate <sup>b</sup> | M | R.Q. | (44) |
| **Hippuric Acids** |  |  |  |
| hippuric Acid | M | R.Q. | (44) |
| **Chlorogenic acids** |  |  |  |
| chlorogenic acid | C | 9.43 (0.57-26.82) | (9) |
|  | T | 17.72 (2.39-40.02) |  |
|  | M | 1.03 (0.01-2.61) |  |

C- Colostrum, T- transitional milk, M- Mature Milk, R.Q.- Relative quantification
<sup>a</sup> The concentrations listed are the highest concentrations reported in each study or a range when more than one concentration is presented. Studies reporting only relative quantification are referred as R.Q (relative quantification).<sup>b</sup> The concentration reflects the sum of two possible isomers;
<sup>c</sup> Highest reported concentration between the three experimental groups in the study (low, medium, and high (poly)phenol consumers);
<sup>d</sup> Converted from µg/L to nmol/L;
<sup>e</sup> The original manuscript states two sulfate isomers for “5-(3-methoxy-4-hydroxyphenyl)-γ-valerolactone-sulfate”. However, this molecule only has one hydroxyl group available for sulfation. For this reason, it is possible that the isomers mentioned in the original manuscript are 5-(3-methoxyphenyl)-γ-valerolactone-4-sulfate and 5-(4-methoxyphenyl)-γ-valerolactone-3-sulfate.

Among the flavan-3-ols, commonly found in chocolate(49), epigallocatechin-gallate was the molecule detected at highest concentrations in breast milk, ranging from 0.67 to 1.12µM (Figure 5A, Table 3). In previously reported intervention studies, reviewed in (50), this molecule was detected in plasma, on average, at much lower concentrations, ranging from 0.03 to 0.38µM(50). Meanwhile, a more recent review shown higher plasma concentrations with a mean Cmax across several studies of 0.87 ± 1.33µM, on par with breast milk concentrations(51). By contrast, other flavan-3-ols like (epi)catechin, was detected at higher concentration in plasma ranging from 0.09 to 1.10µM, in comparison to breast milk of 0.03 to 0.24µM(50). Meanwhile, quercetin was the flavonol detected in highest concentrations in human breast milk ranging from 0.05 to 0.15µM (Figure 5B, Table 3). Remarkably in the study performed by Zhang et al. 2018 measuring quercetin in both breast milk and plasma of the mothers, it was shown that quercetin was more abundant in breast milk and urine in comparison with plasma(44).

**Figure 5.**
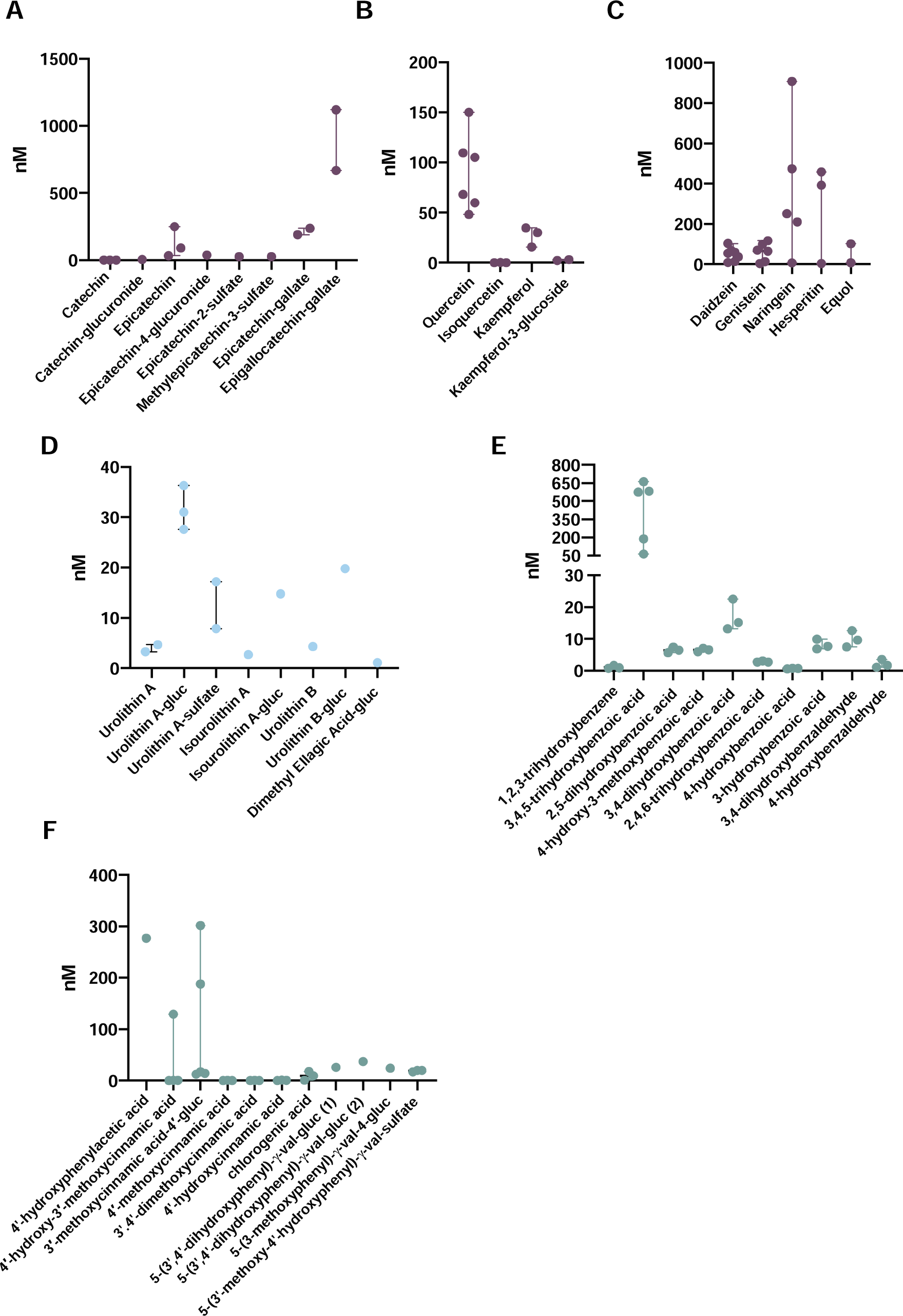
Range of concentrations of (poly)phenols detected in human breast milk. (A) flavan-3-ols, (B) flavonols, (C) flavanones and isoflavones, (D) ellagic acid metabolites, (E) benzoic acids and benzaldehydes, (F) phenylacetic, cinnamic acids, chlorogenic acids and phenyl-y-valerolactones. Each dot represents the different concentrations reported in Tables 3, 4 and 5. Val – valeric acid; gluc – glucuronide.

Regarding the flavanones naringenin and hesperidin, commonly found in citric fruits(49), have been detected in plasma at concentrations ranging from 0.13 to 1.50µM and 0.21 to 0.87µM respectively (Figure 5C, Table 3). Meanwhile, in breast milk both flavanones have been detected at lower concentrations ranging between 0.01 to 0.91µM for naringenin and from 0.004 to 0.46µM for hesperidin (Table 3). Regarding isoflavones, daidzein and genistein have been found in plasma ranging from 0.76 to 3.00µM and 1.26 to 4.50µM, respectively(50). Meanwhile, in breast milk these two isoflavonoids have been found in much lower concentrations ranging from 0.009 to 0.10µM and 0.005 to 0.12µM respectively (Figure 5C, Table 3).

The urolithin metabolites urolithin A-glucuronide and urolithin B-glucuronide were the two most abundant ellagic acid metabolites found in breast milk at 0.03µM and 0.02µM respectively (Figure 5D, Table 4). In plasma these two metabolites have been found at identical concentrations of 0.02µM, similar to those of breast milk(52). On the opposite direction, urolithin A-sulfate detected in breast milk ranging from 0.008 to 0.02µM (Figure 5, Table 4), was detected at much higher concentrations in plasma of 0.45µM(52).

Meanwhile, a recent literature review revealed the full list of low molecular weight phenolic metabolites present in human blood(26). From this list of 137 phenolic metabolites detected in human blood, only 58 have been detected and or quantified in human breast milk (Table 5). Most of these metabolites have been found in significantly higher concentrations in human plasma in comparison with human breast milk. For example, regarding the phenolic acid 3,4-dihydroxybenzoic acid and benzaldehyde 3,4-dihydroxybenzaldehyde, these have been found in breast milk at concentrations ranging from 0.01 to 0.02µM and 0.009 to 0.01µM (Figure 5E, Table 5). While in plasma, these metabolites have been found up to 16µM and 0.03µM respectively. Meanwhile, 4-hydroxyphenyl acetic acid, 4-hydroxy-3-methoxycinnamic acid (ferulic acid) and 3-methoxycinnamic acid-4-glucuronide (ferulic acid-glucuronide) have been found in breast milk up to 0.27µM, 0.13µM and 0.3µM respectively (Figure 5F and Table 5). Whereas in plasma these have been found at 2.6µM, 0.42µM and 0.36µM respectively. Noteworthy, the metabolite detected at higher concentrations in human breast milk was 3,4,5-trihydroxybenzoic acid with a range of 0.06 to 0.66 µM (Figure 5E, Table 5) detected to a lesser extent in plasma 0.12 ± 0.032µM, reviewed in (26).

Interestingly several classes of low molecular weight phenolics have not been detected in human breast milk. This may be related with the lack of standards and/or a target analysis excluding quantification of several metabolites, or the complete absence of this metabolites in human breast milk. Examples of such metabolites not detected in human breast milk are: 2-(phenyl)propanoic acid, mandelic acid, cinnamoylglycine, 4-hydroxy-5-(phenyl)valeric acid, 5-(phenyl)valeric acid, (phenyl)ethanol or 4-ethylbenzenes. Regarding ellagic acid metabolites, besides the 9 ellagic acid metabolites found in human breast milk and found in blood, others have been found in human blood but not in human breast milk, namely urolithin C, urolithin C glucuronide and urolithin C sulfate (34).

Overall, several reasons might explain the discrepancy between the number of (poly)phenols detected in blood versus human breast milk. Noteworthy the data obtained for plasma and breast milk samples have been obtained from different populations with probably differences in the dietary exposure of (poly)phenols and in the absorption and metabolism of these compounds. A particular relevant question is the possible changes on the metabolism of (poly)phenols of recent post-partum volunteer. Large studies quantifying (poly)phenols in both plasma and breast milk of the same volunteer are required comparing concentrations in both fluids and addressing this possibility. In the paper of Zhang *et al.*, most of the identified metabolites were detected in the three biological samples (plasma and urine of two volunteers and breast milk of another volunteer) and some were present in urine and breast milk but not in plasma. This may reflect some intrinsic physical chemical property of each compound to either stay in plasma or being rapidly eliminated from blood and accumulate in urine and breast milk. Other factors could be i) their absence from human breast milk due to the lack of specific transporters impairing their path through the mammary tissue, ii) the timepoints used for the sampling and further quantification of the (poly)phenols metabolites in the milk iii) lower concentrations in human breast milk (below limit of quantification) in comparison with the already reported in circulation, iv) the fact these (poly)phenols metabolites have never been searched in human breast milk due to the lack of available standards, v) the extraction method is not fully compatible to all (poly)phenol metabolites.

One unsolved question that its understanding can shed light to some of these questions is the mechanism of transport allowing (poly)phenols and/or metabolites to reach human breast milk. The transport and delivery of (poly)phenols to the mammary glands is still unknown and may result from passive diffusion, active transport, or carrier mediated transport. For flavonoids, several methods of transport have been proposed to cross several tissues like the gut from passive diffusion to mediated transport, especially by ATP-binding cassette transporter (ABC transporters) and organic acid transporters (OAT), reviewed in (53). For phenolic metabolites it is hypothesized that passive transport might be the main route of transport across the tissue, although some transporters have already been described. These include gallic acid through OAT3 (54) and homovanillic acid through rOAT3 (55). Interestingly homovanillic acid is both an endogenous molecule, a metabolite of dopamine and a phenolic metabolite result of microbiota catabolism of flavonoids. Other phenolic metabolites that also result from endogenous pathways like hippuric acid and 2-(3,4-dihydroxyphenyl)acetic acid (DOPAC) that might benefit from the endogenous routes of transport. Furthermore, it also offers the possibility that structurally similar phenolics might share the same routes.

## 4 The potential health benefits of (poly)phenols and phenolic metabolites in human breast milk

Although multiple studies have reported the protective effects of (poly)phenols against inflammation, oxidative stress and cardiometabolic stress, and the potential of isoflavones as phytoestrogens, the full spectrum of effects of (poly)phenols in human breast milk, both from the mother and the newborn perspective, has not been fully explored. Across the different studies reported in the literature the evaluation of the impact of (poly)phenols detected in human breast milk have been rather limited to the anti-oxidative properties of milk or the quantification of phytoestrogens and its impact on the newborn development.

Regarding the anti-oxidative properties of (poly)phenols in human breast milk Poniedzialek *et al.*, showed that an increase in total (poly)phenols in human breast milk associated with a higher total antioxidant capacity that were positively correlated with vegetable consumption (56). Total (poly)phenols also correlated negatively with malondialdehyde content. Noteworthy it is important to bring attention to the high degree of variability of samples collected. Li et al., compared the total (poly)phenol content in samples from breast milk and infant formula. In this case no significant change was detected as the variability within breast milk and infant formulas was significant (422 – 751 mg/kg and 329 – 797 mg/kg of ferulic acid equivalents respectively) (57).

A different study by Silberstein *et al.*, showed that the (poly)phenolic content in breast milk seems to be influenced by maternal health status (58). The study has shown that the breast milk composition of preeclampsia women presents a 33% increase in total (poly)phenol content in comparison with healthy women (58). The increase in total (poly)phenols in mothers with preeclampsia was associated with a reduction in about 20% the levels of lipid oxidation in colostrum in comparison with healthy women (58). Unfortunately, the authors only measured the total amount of (poly)phenols and did not investigate the presence or effects of specific (poly)phenols or their metabolites.

Concerning isoflavones, these have been classified as phytoestrogen for their chemical similarity with estradiol and the capability to induce estrogenic or anti-estrogenic effects depending on the dose (59). Isoflavones as phytoestrogens bind to estrogen receptors alpha and beta (59). Isoflavones are present across several fruits and legumes with significant relevance in soybeans and soy-based produces like soy milk and tofu. For their capability to act as phytoestrogens, soy and soy-based products have been used in infant formulas in the last century and their effects analyzed.

Setchell et al., compared the presence of isoflavones in 4-month-old infant blood samples consuming human breast milk, soy-based infant formulas, and cow-based milk formulas (60). The concentration of isoflavones circulating in babies fed with human breast milk was on average 4.7 ug/L in comparison with the 9.4 ug/L of cow-based infant formula and the 979.7 ug/L of soy-based formula. This high plasma isoflavones concentrations in infants consuming soy-based formula are however much higher than the normal estradiol concentrations for infants of this age. One phenomenon detected was the lack of isoflavones metabolites, namely equol and its metabolites, from soy-based infant formula since the formation of this metabolites are largely dependent on the gut microbiota of the mother (60).

Meanwhile, Rosa et al. also showed a significant difference on the urinary phenolic metabolites of 3-month-old infants feed with breast milk, cow-based infant formula, and soy-based infant formula (61). In this study the overall quantity of phenolic metabolites excreted in infant urine was 2-fold higher for some metabolites, namely 3′-hydroxyphenylacetic acid and 4′-hydroxyphenylacetic acid in infants feed with soy-based infant formula in comparison with human breast milk feed infants (61).

Although the evidence suggests an increase of isoflavones levels in infants fed with soy-based formulas there is no clear evidence of a health benefit. In fact, a meta-analysis could not find any negative effects of a high isoflavone intake from soy formulas, but also no benefits, on endocrine and other physiological functions of breastfeeding (62). The authors speculated that most isoflavones detectable in the plasma of soy fed infants are conjugated to sugars, which cannot link to isoflavone receptors.

Regarding the cognitive developmental status of infants, Andres *et al.*, 2012 evaluated 1 year old infants consuming breast milk, cow-based milk formula or soy-based milk formula. In this study infants were assessed using Bayley Scales of Infant Development (BSID), from which the Mental Developmental Index (MDI) and Psychomotor Development Index (PDI) were derived (63). Small but statistically significant effects of the diet on the MDI scores were seen for infants 6 months or older. Breastfed infants had significantly higher scores than soy fed infants at ages 6, 9, and 12 months and significantly higher scores than cow-based milk fed infants at ages 9 and 12 months. Although these effects reached statistical significance, the MDI scores were within the expected normal range, and the differences were very small. Significant effects of the diet on the PDI scores were transient. Soy fed infants had significantly lower PDI scores, although small, compared with breastfed infants, and seen only at 6 months. Furthermore, infants were also assessed with the Preschool Language Scale-3 (PLS-3) by using the expressive communication and auditory comprehension subscales. In PLS-3, only cow-based milk fed infants had significantly lower scores compared with breastfed infants at ages 3 and 6 months, but again all groups were inside the normal range (63).

Ostrom *et al.*, also observed the immune status of infants fed with soy-based formulas and human milk/infant formula feed infants (64,65). In this study infants were immunized against *Haemophilus influenzae* type b polysaccharide, diphtheria toxoid, tetanus toxoid, and oral poliovirus of children and their responses evaluated at 6, 7 and 12 months of age. Antibodies against *H. influenzae* type b at 7 and 12 months were higher significantly higher in infants fed with soy-based milk (65). In contrast, human milk/infant formula fed infants had higher poliovirus neutralizing antibodies at 12 months of age. Serum IgG and IgA levels were similar between groups as well as morbidly through the first year of life. Only physician-reported diarrhea was different between groups fed with human milk/infant formula having less cases than the soy group (65). Latter Cordle et al., with the same cohort showed that the number and percentage of B, T and NK lymphocytes were within age-related normal ranges with minor differences between soy formula and human milk/infant formula fed infants (64). Overall, no differences were detected between the two groups due to soy-based formulas consumption suggesting normal immune system development.

## 5. Conclusions and future perspectives

In this literature search we have listed the (poly)phenols and their metabolites, namely flavonoids, ellagic acids and phenolics detected in human breast milk. Nonetheless, the information available in the literature about the presence of (poly)phenols and/or their metabolites reported in human breast milk is still in its infancy and requires further studies. Many (poly)phenols and their metabolites have been detected in human circulation (26), yet their presence in human milk have not been fully explored, due to several factors mentioned before. Meanwhile, several nuances in the design of the studies performed may be hindering the identification and their quantification in breast milk such as the timepoint of collection, volume, extraction and method of quantification that may still require further optimizations.

Concentrations of some (poly)phenol and their metabolites in human breast milk may be dependent on the diet of the mother, demonstrated by some intervention studies using (poly)phenols rich foods or the stratification of volunteers. This type of pattern has also been observed, with an increase in sugar and fat in human breast milk after mother ingested sugar and fat enriched diets (66). The type of milk (colostrum, transitional or mature) did not seem to make an impact in the quantification of some (poly)phenols. Several factors may still require further investigation such as mother’s interindividual differences (genetic, metabolic, microbiota) or even variations caused by the circadian rhythm on breast milk composition (66). In fact, most studies have not reported the time of collection and volume of breast milk, which may be a big limitation, as milk production and composition may vary according to the mothers and child dietary needs across the day. Some of the mentioned studies have managed this issue by assuring the collection of the full content of breast milk, choosing a specific time of the day for collection of all volunteers. Notwithstanding, information on how (poly)phenol composition varies on breast milk during the day is still nonexistent. Additionally, most studies have used small sample sizes that may overlook interindividual differences. Further studies addressing these issues may give a better insight on the presence of (poly)phenols and/or their metabolites in human breast milk and the impact of diets like the Mediterranean diet, vegetarian diet or vegan diets on the compounds present in human milk.

The studies on the potential health benefits to the infant of mother’s (poly)phenols consumption and their presence in breast milk have been limited. The anti-oxidative properties of breast milk as function of total (poly)phenols have been performed, yet these should be looked has rather rough indicators. Measurement of general markers of antioxidant status, such as ORAC, TRAP, FRAP and TEAC are inappropriate methods for clinical studies because of the broad range of compounds (e.g., uric acid, albumin) that may interfere with the results of these analyses (67,68). More suitable markers for the increased cellular antioxidant potential may include nuclear Nrf-2 DNA binding capacity and NQO1 protein levels, which were observed to increase following (poly)phenol intake and not following a challenge meal alone (69).

The presence of phytoestrogens like isoflavones in human breast milk has been studied due to their potential to mimic estrogens. Several studies have focused on the comparison between human breast milk and soy-based infant formulas rich in isoflavones. Nonetheless, no clear conclusion has been reached about the potential health benefit of soy-based milk. Although (poly)phenols have been known for their anti-inflammatory properties, until now only one study has focused on the immunity of the infant and the presence of (poly)phenols in the breast milk (64). Newborns and especially pre-term infants can suffer from severe inflammatory conditions yet our knowledge of (poly)phenols impact in these conditions is lacking as this study has never been addressed.Overall, there are clear gaps in our current knowledge concerning what metabolites from dietary (poly)phenols appear in human breast milk and their quantification. Moreover, this hampers our understanding of their potential health benefits for the infant. At the present date the scientific data on the health potential of (poly)phenols in adults is vast, yet very little is known about their effect in infants that received them through milk. It will be important to design studies that make it possible to relate the milk composition in (poly)phenol metabolites with infant’s health outcomes. Our understanding of the potential health benefits of mother’s diets rich in (poly)phenols for the newborn have a huge potential to help clinicians to deal with such a fragile phase of life of infants with health complications. This will open a new way for the mothers to provide the conditions for a strong and healthy growth of their child.

## Data Availability

All data reviewed in this manuscript are contained within the cited studies

## Abbreviations

BM: breast milk
BSID: Bayley Scales of Infant Development
DOPAC: 2-(3,4-dihydroxyphenyl)acetic acid
EHC: enterohepatic circulation
FRAP: ferric reducing antioxidant power
LMW: low molecular weight
MDI: Mental Developmental Index
OAT: organic acid transporter
ORAC: oxygen radical absorbance capacity
PDI: Psychomotor Development Index
PLS-3: Preschool Language Scale-3
SC: systemic circulation
SPE: solid phase extraction
TEAC: trolox equivalent antioxidant capacity
TRAP: total reactive antioxidant potential

## Acknowledgments

The authors gratefully acknowledge the support given by Doctor Teresa Costa regarding the literature search process. This work has received funding from the European Research Council (ERC) under the European Union’s Horizon 2020 research and innovation program under grant agreement No 804229. INOVA4Health Research Unit (LISBOA-01-0145-FEDER-007344). Authors would also like to acknowledge FCT for financial support of D.C. (2020.04630.BD).

## 6. Author contributions

**Diogo Carregosa**: Conceptualization; Data curation; Formal analysis; Investigation; Methodology; Resources; Software; Visualization; Writing – original draft. **Inês P. Silva**: Data curation; Formal analysis; Investigation; Methodology; Resources; Software; Visualization; Writing – original draft. **Carolina Teixeira**: Data curation; Formal analysis; Investigation; Methodology; Resources; Software; Visualization; Writing – original draft. **Mariana Baltazar**: Data curation; Formal analysis; Investigation; Methodology; Resources; Software; Visualization; Writing – original draft. **Rocio García-Villalba**: Formal analysis; Supervision; Validation; Visualization; Writing – original draft; Writing – review & editing. **Filipa Soares Vieira**: Validation; Visualization; Writing – review & editing. **Mónica Marçal**: Validation; Visualization; Writing – review & editing. **Madalena Tuna**: Formal analysis; Supervision; Validation; Visualization; Writing – original draft; Writing – review & editing. **Cláudia Nunes dos Santos**: Conceptualization; Data curation; Formal analysis; Project administration; Supervision; Writing – review & editing

